# Trends in Perception of COVID-19 in Polish Internet

**DOI:** 10.1101/2020.05.04.20090993

**Authors:** Andrzej Jarynowski, Monika Wójta-Kempa, Vitaly Belik

## Abstract

**INTRODUCTION:** Due to the spread of SARS CoV-2 virus responsible for COVID-19 disease, there is an urgent need to analyse COVID-2019 epidemic perception in Poland. This study aims to investigate social perception of coronavirus in the Internet media during the epidemic. It is a signal report highlighting the main issues in public perception and medical commutation in real time.

**METHODS:** We study the perception of COVID-2019 epidemic in Polish society using quantitative analysis of its digital footprints on the Internet on platforms: Google, Twitter, YouTube, Wikipedia and electronic media represented by Event Registry, from January 2020 to 29.04.2020 (before and after official introduction to Poland on 04.03.20). We present trend analysis with a support of natural language processing techniques.

**RESULTS:** We identified seven temporal major clusters of interest on the topic COVID-2019: 1) Chinese, 2) Italian, 3) Waiting, 4) Mitigations, 5) Social distancing and Lockdown, 6) Anti-crisis shield, 7) Restrictions releasing. There was an exponential increase of interest when the Polish government “declared war against disease” around 11/12.03.20 with a massive mitigation program. Later on, there was a decay in interest with additional phases related to social distancing and an anti-crisis legislation act with local peaks. We have found that declarations of mitigation strategies by the Polish prime minister or the minister of health gathered the highest attention of Internet users. So enacted or in force events do not affect interest to such extent. Traditional news agencies were ahead of social media (mainly Twitter) in dissemination of information. We have observed very weak or even negative correlations between a colloquial searching term ‘antiviral mask’ in Google, encyclopaedic definition in Wikipedia “SARS-CoV-2” as well official incidence series, implying different mechanisms governing the search for knowledge, panic related behaviour and actual risk of acquiring infection.

**CONCLUSIONS:** Traditional and social media do not only reflect reality, but also create it. Risk perception in Poland is unrelated to actual physical risk of acquiring COVID-19. As traditional media are ahead of social media in time, we advise to choose traditional news media for a quick dissemination of information, however for a greater impact, social media should be used. Otherwise public information campaigns might have less impact on society than expected.

## INTRODUCTION

Although a large part of the Polish population has heard about coronaviruses for the first time a few weeks ago, in reality they face less dangerous coronaviruses causing simple cold all the time. Only the emergence of a novel strain – SARS-CoV-2 from Wuhan gave the word “Coronavirus” (pl “Koronawirus”) a new dramatic meaning. Before disease introduction, less than a half of surveyed Poles believed that “Coronavirus” is the most important topic in the second half of February 2020 (1). The disease was not detected in Poland until 04.03.2020. This topic did not receive much attention until the introduction and only started to drive media and social life afterwards. During opinion polls performed on 09-10.03.20, 63% of respondents reported that this is a serious threat for Poland, 40% that this is a serious threat for their family and 73% that this is a serious threat for the Polish Economy (2). Opinion polls performed on 25-27.03.20 showed that almost 90% surveyed Poles declared “Koronawirus” as the most important current threat for them (3) [to disclose - different methodologies were used in each of these studies].

Risk perception is not as straightforward as the term used in risk assessment, i.e. value at risk - expected value of loss (infection or death) with given probability of an event, but a complicated socio-psychological construct with biases and errors (4). For instance, on average people believe that negative events such as infection and death are less likely to happen to them than to others (5). In the previous epidemics (A/H1N1, SARS, Ebola) in the Internet era societies (such as Polish (6)) experienced quite similar phases of interest starting from fear, then moving to practical issues such as prevention and finishing with scapegoating (7). Recently media activities are being analysed worldwide to better understand perception and spread of diseases (8) as COVID-19 (9). Social media can serve as a valuable source of information as well as disinformation about the virus globally, fuelling panic and creating so-called infodemics (10), at unprecedented speed. Propaganda and persuasion techniques are widely used on the Internet, easily reaching certain target groups susceptible to conspiracy theories and effectively polarizing societies (9, 11). Panic-related behaviours accompanying a virus outbreak (7) are influencing the epidemic spread, with the Internet being the main mediating mechanism. This could lead to destructive collective behaviour, such as e.g. xenophobia against people from affected countries (12). Moreover, infodemiology and infoveillance (10) should be employed for standard surveillance purposes within the emerging field of “digital epidemiology” and in the area of public health.

In this study, by quantitative analysis of “digital traces/footprints” on the Internet (social and traditional media), we concentrate on the number and nature of events such as information queries and information propagation. The Internet, social media, apps and web-information are now our every-day life experience and practice. Tracking “digital footprints” of Internet users brings understanding of customers’ behavior and preferences on a large scale in the commercial and scientific sectors. Up to our knowledge there were no previous studies quantitatively linking the Internet activities and perception of infectious diseases in Poland (13, 14), except for our own studies (15, 16) and some straightforward attempts such as correlating the incidence of influenza with Google Trends (8). Moreover, during the first few weeks of COVID-19 pandemic a tremendous amount of studies appeared targeting English and Asian languages (17) with an underrepresentation of European countries. Thus the present study is a first exploratory attempt filling this gap and continuing a preliminary research on “Koronawirus” perception in Poland before disease introduction (15) representing a more general approach highlighting the role of social sciences during COVID-19 pandemic (16) in public health.

The aim of this article is to present some common tools useful in studying epidemic dynamics. We collected the information-seeking behaviors (digital paths) and compared them with real time events. These efforts are our contribution to public health surveillance (17) by collecting and analyzing health-related information. We argue that presented tools are able to track population’s behavior and monitor or even predict (local) outbreaks of diseases or abnormal behaviors (like social panic or harmful beliefs caused by fake news). Such data analytics may provide near real time outbreak information in various formats, independently from official statistics or surveys reports. We aimed to describe phases of interest of SARS-CoV-2 in Polish Internet, as well as to indicate the most important topics of social perception of the new disease.

## METHODOLOGY

We primarily analyzed quantitative digital footprint data on the Internet from January 2020 to 29.04.2020, including their representativeness. The unit of our analysis is a daily interest, defined in a different form for each platform. As a main keyword in investigated platforms (14) we chose a colloquial term “Koronawirus”/”Coronavius” due to its high penetration in the society. Other related keywords in use are much less popular, except for Wikipedia, where the medical term SARS-CoV-2 was frequently chosen (16). For comparative perspective we collect data from the Polish Internet to archive the highest possible range of communication types and channels:

a. from Google a daily relative value of “Koronawirus” in terms of queries intensity and news picked by Google from Google Trends;
b. from YouTube a daily relative value of “Koronawirus” in terms of queries intensity from Google Trends;
c. from Twitter a daily number of Tweets with #Koronawirus hashtag using the Twitter API (the only one allowing us to measuring the active Internet usage - production and retweeting of information);
d. from EventRegistry a daily number of news/articles with the topic “Koronawirus”;
e. from Wikipedia a daily number of page visits for “SARS-CoV-2” articles.

It is important to underline that data can be biased e.g. due to content presenting algorithms on the media platforms. For instance, technological giants Google and Twitter are supposed to implement fact verification algorithms to filter out false information and promote official information (e.g. showing widgets related to COVID-19) and we can rely only on their searching engines after filtration. This may jeopardize the reproducibility of our results. In our study each considered Internet platform is described separately and has its own target audience and communication style. The main data comes from Google Trends due to the highest share of Polish Internet users. Despite some disadvantages and exploratory nature of this study, it provides us with an opportunity to analyse a huge amount of digital footprint data at low cost and in a short time.

Our research includes further methodological limitations. First, the population structure of the Internet users may lead to under/over-estimation of observed phenomena. We cover ca. 28 millions Polish language speaking residents of Poland representing over 85% of the literate population (18). The Polish Internet users are younger (especially among content creators (19)) with females prevailing (especially among content receivers (20)) as compared to the average population. Our results do not describe the perception of the oldest groups, due to digital exclusions, health literacy limitations or consideration of social media as less informative (21). Moreover, we have not studied the geographical or social status variability. Although the collected data can be successfully treated as a “digital proof”, the research on this phenomena should be considered *in statu nascendi,* therefore the conclusions of our analysis should be verified by in-depth triangulational studies (22).

## RESULTS: GOOGLE TRENDS

Google has 95% share among Polish Internet users (around 26 Millions) with over 8 billion entries per month and is the undisputed leader on the Polish Internet market (18). Interest in novel Coronavirus on Google can be measured by the number of search queries (Fig. 1). According to own calculations based on Google Ads there were around 5 million searches of term “Koronawirus” in March 2020 (Google does not provide absolute search values). YouTube has a 68% share among Polish Internet users with about 700 million entries per month (18). In addition, streams from the mobile app should be taken into account, as it is the most popular app on Poles’ smartphones (18). As YouTube belongs to Google corporation, it is a part of Google Trends solution and for our analysis, we selected videos on the main subject of "Koronawirus".

**Figure 1.**
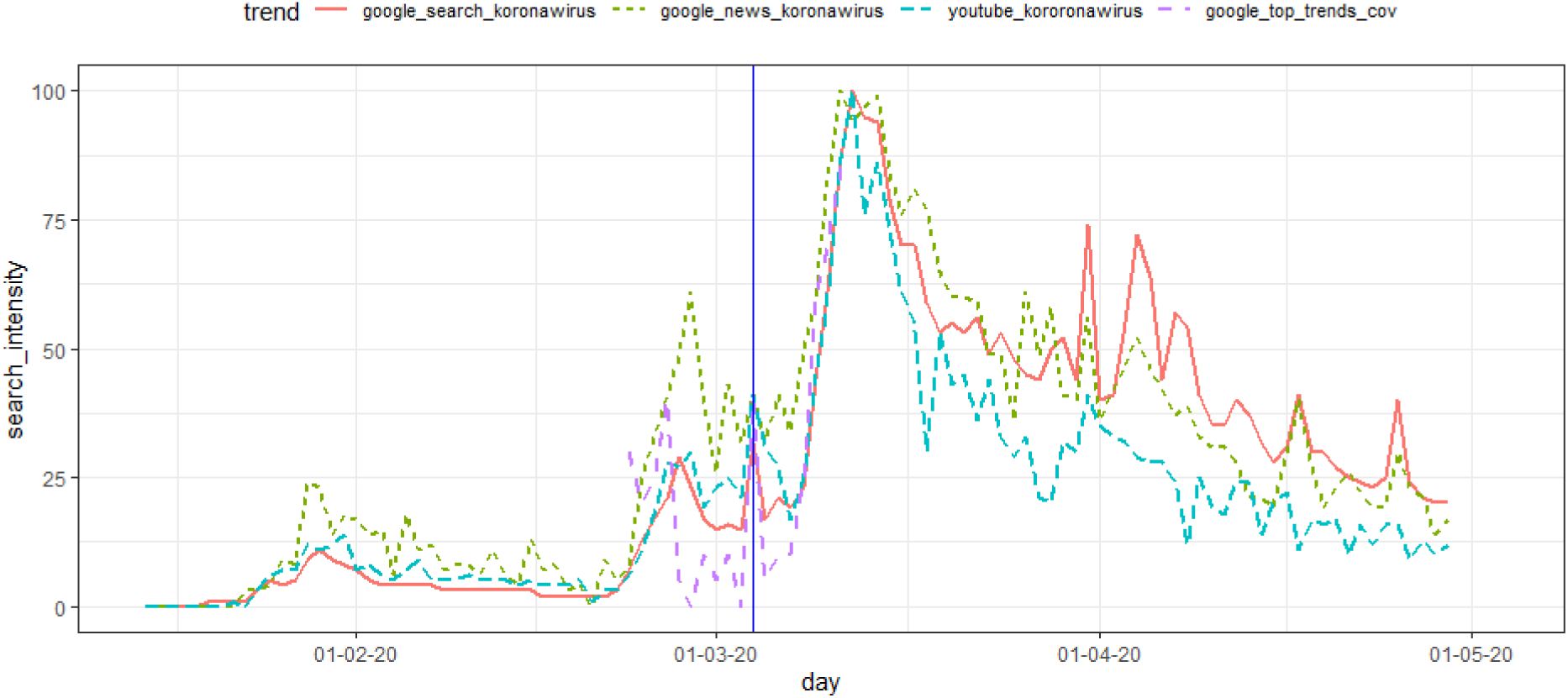
The intensity of searched queries and news with the word “Koronawirus” in Polish Google and searched queries with the word “Koronawirus” in Polish YouTube (15.01-29.04.20) and percentage of “Koronawirus” related queries in Google Top Trend Topics (23.01-11.03.20) both generated using Google Trend tool. Disease introduction in Poland is marked with the vertical line.

The topic of the “Koronawirus” occurred in Polish Internet first time in the middle of January. Before the disease introduction to Poland three phases of interest can be distinguished. They were analyzed in our former publications (15, 16). The next three stages may be considered after the outbreak (Fig. 1):

1. China-related phase: from the end of January and beginning of February when the epidemic was announced and confirmed in China. We observe a small peak around 25.01.20 (e.g. death of Liang Wudong) and around 29.01.20 (e.g. first case in Germany);
2. Italy-related phase: from the middle of February till the end of February (when the number of infections rapidly increased in Italy). We see a clear peak around 27.02.20 (e.g. fake news about possible introduction of the disease to Poland (23));
3. Waiting phase: beginning of March (waiting for introduction - decreased attention). We see a small peak on introduction day (04.03.20);
4. Mitigation phase: around 11/12.03 - mitigation (declaration of main restrictions) and epidemic (real-time R_0_~2.5 estimation) phase with the highest interest in “Koronawims”;
5. middle of March - “social distancing” and “lockdown” phase (real-time R_0_~2 estimation) with small peaks corresponding to new restriction;
6. from the end of March and the beginning of April - anticrisis act (in Polish media called “Anticrisis shield”) phase with high peaks corresponding to anticrisis shield declarations (real-time R_0_~1.5 estimation).
7. Second part of April. Restrictions releasing phase (real-time R_0_~1.2 estimation).

It is important to note, that until disease introduction to Poland there were no Coronavirus related searches in top 25 Google queries at all. Although “Koronawirus” related queries were observed in top trends before the introduction of the disease to Poland, they start to dominate top trends queries only after the massive mitigation measures were taken such as school/university and border closures declarations between 09-12.03.20 (Fig. 1). After the first confirmed case in the country we can see a peak on that day (04.03.20) and substantial growth after important measures were implemented by Polish authorities (09-12.03.20).

**Figure 2.**
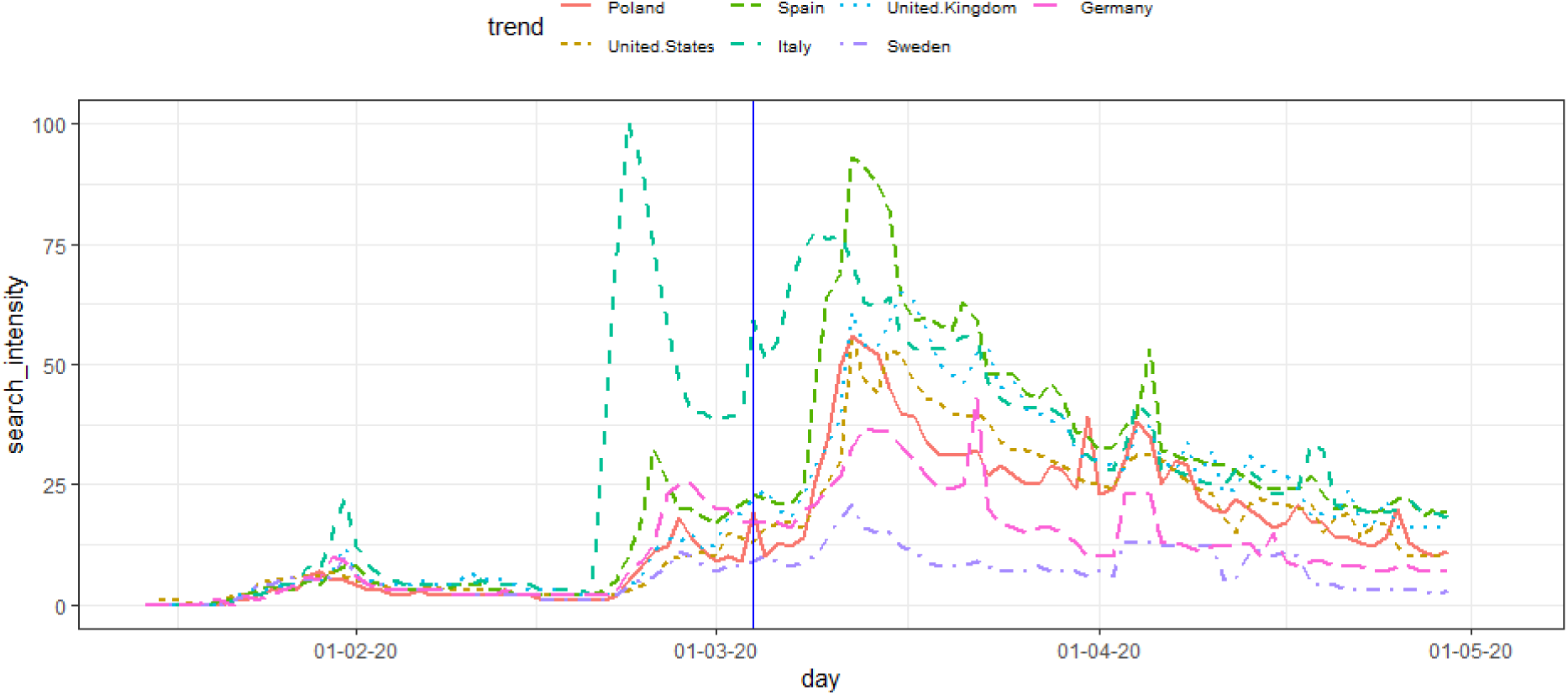
The intensity of the topic “Coronavirus” in Google for Poland, Italy, Spain, USA, UK, Germany and Sweden (15.01-29.04.20) generated using the Google Trend tool. Disease introduction in Poland is marked with the vertical line.

Search activity for information on the infection in Poland was still much lower than in countries with higher global connectivity (24), which earlier became epidemic (Fig. 2) with some exceptions such as Sweden (a country with a significantly different approach towards COVID-19 mitigation). On the last day of the investigated period – 07.04 – there were 4848 officially laboratory confirmed cumulative cases, so official prevalence in Poland was around 100 per million inhabitants, which was below the European Union average.

## RESULTS: OTHER PLATFORMS

Wikipedia traffic is another indicator of social activity. Wikipedia has an Internet coverage of 57% with over 350 million page visits per month among Polish Internet users (18). There is a significant overrepresentation of users with tertiary education inhabiting big cities (affinity index>120 (25)). We looked at the history of page views and discussions around the articles “SARS-CoV-2” (pl.wikipedia.org/wiki/SARS-CoV-2). There were 492584 total page views on the article “SARS-Cov-2” (10.02-07.04.20).

We choose EventRegistry (eventregistry.org) as a traditional media search engine because it has a large range of online news services representing a broad political spectrum. Besides, it gives priority to digital versions of other broadcasting channels, including television, radio or newspapers. During 15.01-07.04.20, 404897 representative articles were selected (the non-systematic sampling method was applied) with inclusion criteria: keyword – “Koronawirus” or topic – Coronavirus in an article body, and country – Poland and language – Polish. In the traditional media the weekly seasonality of articles (less articles during weekends on average) is especially prominent.

Twitter in Poland has relatively low popularity (~3 million registered users or less than 8% of the country population) and is mainly used by expats, journalists and politicians (26). However, Twitter provides an API for data acquisition which allows us to collect 704548 tweets with #Koronawirus hashtag during 15.01-07.04.20.

**Figure 3.**
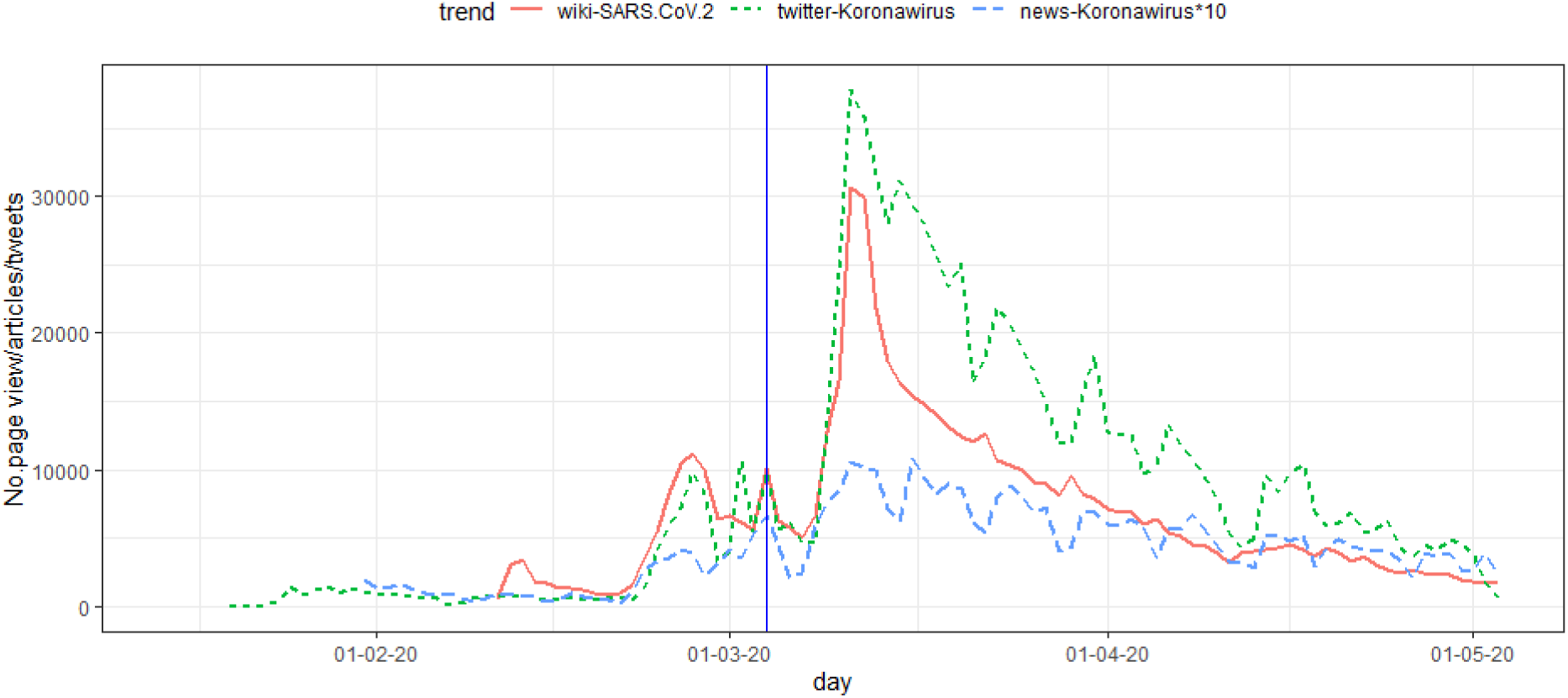
The intensity of topic “Koronawirus” on various media platforms. For Twitter: Number of tweets per day with the Koronawirus hashtag in Polish language (15.01-02.05.20). For Wikipedia: The number of views on the article “SARS-CoV-2” (10.02-02.05.20). For News: Number of articles (31.01-02.05.20) generated using Event Registry (multiplied by 10). Disease introduction is marked by the vertical line.

## DISCUSSION: COMPARISON OF DIFFERENT PLATFORMS

To compare the interest on COVID-19 on different Internet platforms we visualized the available queries and interest measurements as time series together and marked events important to the Polish public (Fig. 4). The largest peak of interest in our analysis for all platforms on 11/12.03.20 was a day of the pandemic declaration by WHO and the declaration of massive mitigation program (27) in Poland. We see that traditional news agencies (well represented in Event Registry) as well as Google searches could precede and form more distinct and sharp peaks of interests than social media platforms. Moreover, discussions on social-content media (YouTube and Wikipedia) demonstrate smoother interest trajectories (Fig. 4) than information providers (news from EventRegistry, Google).

**Figure 4.**
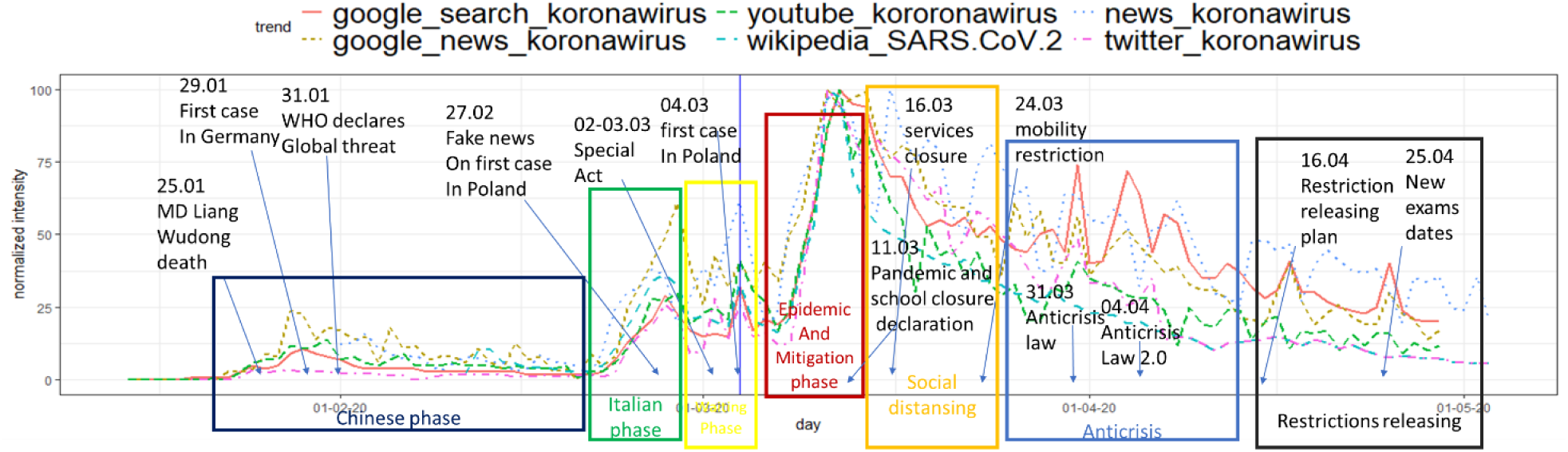
The intensity of the topic “Koronawirus” on various media platforms during 15.01-07.04.20. Time series were normalized to 100 by maximal value for a given series. Six phases of interest: 1) Chinese phase (COVID-19 emerges in Asia (28); 2) Italian phase (second wave of COVID-19 (28)); 3) Waiting phase (awareness building and waiting for a confirmed first case in Poland); 4) Epidemic and Mitigation phase; 5) Social distancing and Lockdown phase; 6) Anticrisis acts; 7) Restrictions releasing. Disease introduction is marked by the vertical line.

We detected time lags between different platforms for given topics (e.g. traditional news media are ahead of commentary media in the case of fake news on a possible disease introduction) in phases 1-4 (Fig. 5). It could imply that social media disseminate information via word-of-mouth spreading mechanisms. On the other hand, traditional media have journalists hired to search and select most interesting topics quickly (29). It leads to faster response to events by news agencies (represented by EventRegistry) than social media. The strongest lag (1 day) is observed between EventRegistry (the most ahead of) and Twitter (the most delayed) (Fig. 5).

**Figure 5.**
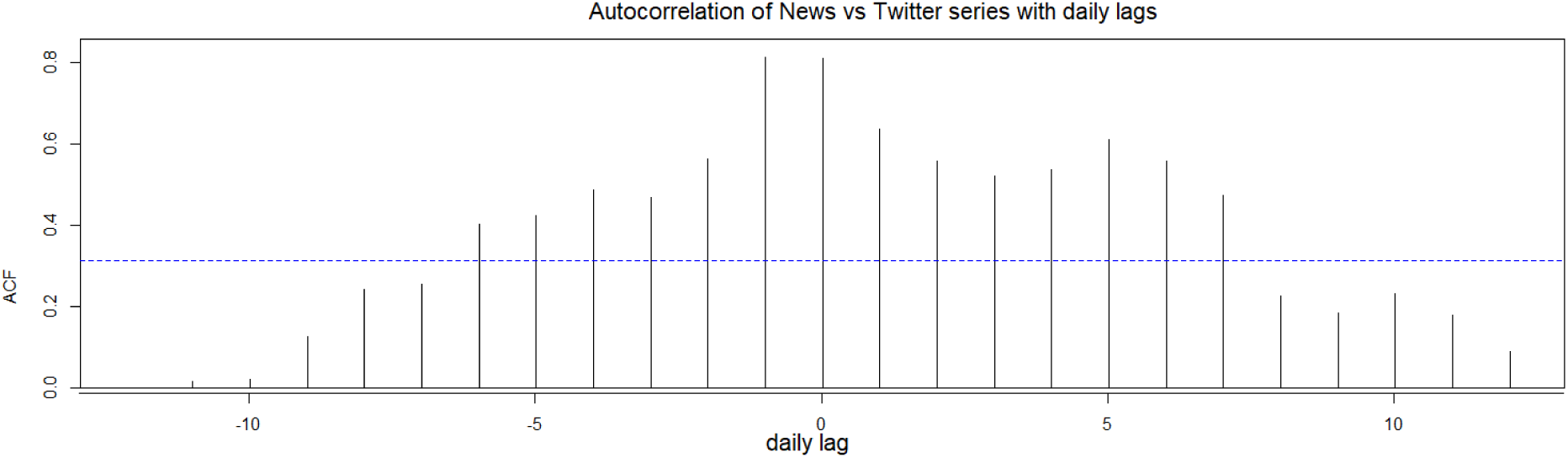
Lagged (in days) correlation between daily series of article counts from EventRegistry (news) and Tweets numbers (31.01–14.03.20) on COVID-19 related topics.

We detected time lags between official SARS-CoV-2 incidence and risk perception. For instance interest in protective masks ahead incidence 3 days before. On the other hand increase in incidence can be linked with decrease of “Koronawirus” search in Google 6 days later [Fig. 6].

**Figure 6.**
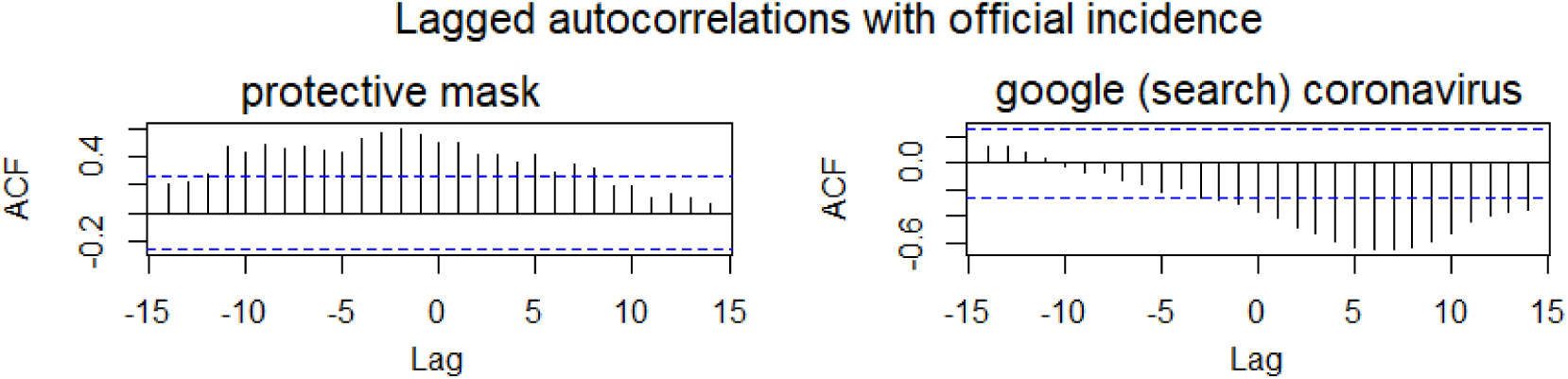
Lagged (in days) correlation between daily series of official SARS-CoV-2 incidence and protective mask and google “Koronawirus” searches (04.03–29.04.20).

To quantify the similarity in interest on different platforms we calculated Pearson correlations coefficients of the time series (Fig. 7). Additionally, the popularity of terms related to COVID-19 were also investigated. Due to limitations imposed by the Editorial Board they are presented in the APPENDIX. The majority of measured terms are positively correlated. *Protective mask* as a signal of fear/perception of risk is less correlated with other variables, more related to information needs. *Protective mask* and *washing hands* pair is much less correlated than *protective mask* and *hand disinfection* pair, which could mean that people search for professional solutions rather than simple and practical ones. Moreover, high amounts of search queries such as *antiviral mask* (there is no such medical term) implies that people are searching protection measures defined in non-professional terms (15). Official COVID-19 incidence correlates only weakly positively or even negatively (Fig. 6) with measured interest value so risk perception in Poland is significantly unrelated to actual physical risk of acquiring infections, which is contradictory to the situation in some other countries (28).

**Figure 7.**
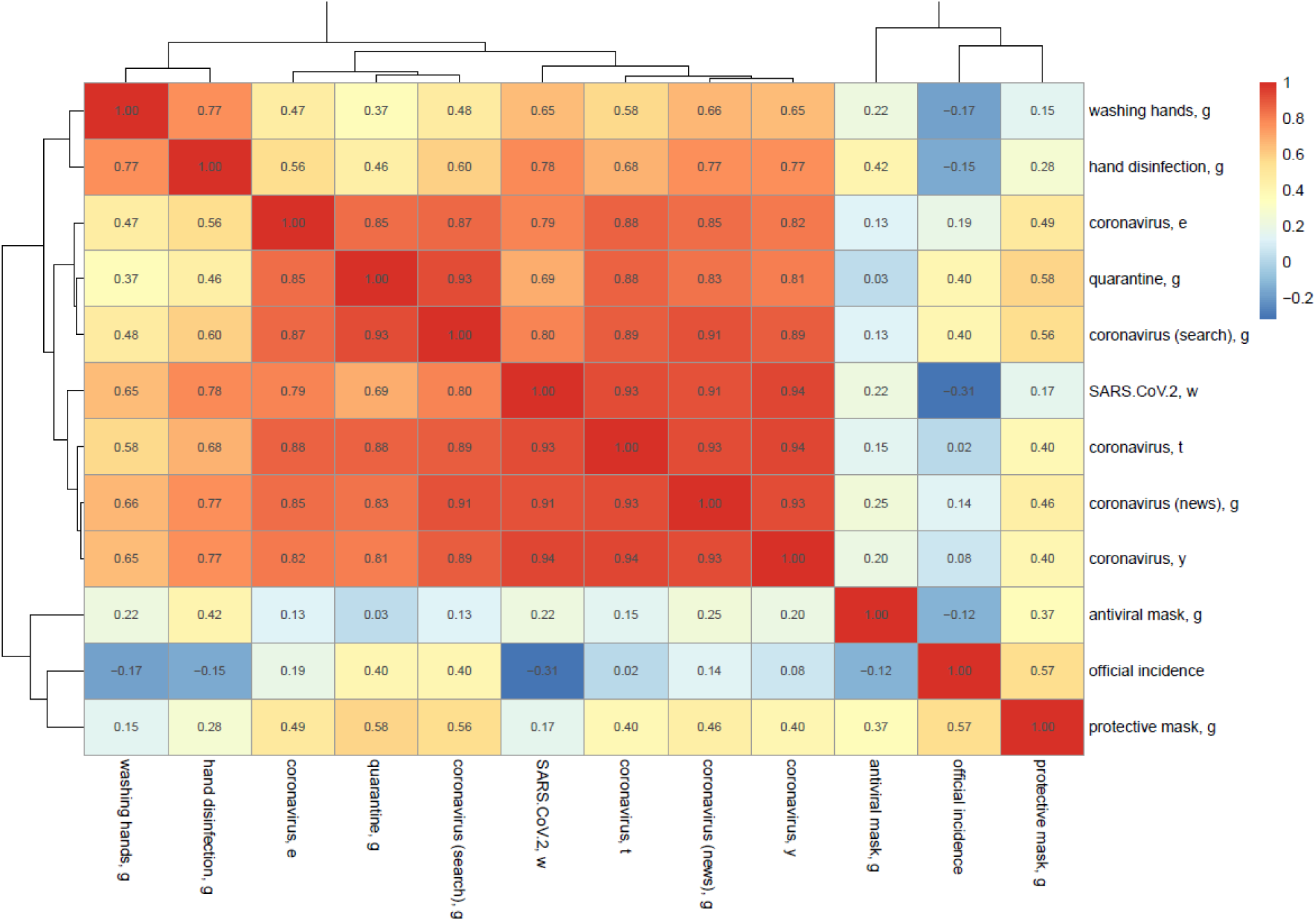
The Pearson’s correlation matrix and the corresponding hierarchical clustering for the terms “Koronawirus”/”Covronavius” and related epidemiological queries on various media platforms for pairwise observations during 15.01-07.04.20 period (g – Google, w – Wikipedia, y – Youtube, t – Twitter, e – EventRegistry) and official COVID-19 incidence. With the significance level.05 all correlations were significant except for correlations related to “antiviral mask”, “official incidence” and the pairs “protective mask, g”/“washing hands, g” and “protective mask, g”/“SARS-CoV-2, w”. Scale corresponds to correlation strength.

## CONCLUSIONS

In the face of the COVID-19 pandemic we have to deal with an unprecedented flood of information (information ’noise’) and our task was to extract important events and features from the broad range of the Internet media in Poland. Observing the amount of the Internet queries we have to state that citizens have treated the Internet as a very important (and we assume available) source of knowledge. Our approach targeted quite a wide range of general population with relatively high coverage of Internet users with quite a significant audience variability across platforms. It’s important to mention that our analysis operated on subjective populational perception and there is no direct translation between scientific evidence (e.g. the question of the effectiveness of protective masks in infection prevention) and colloquial knowledge describing infectious disease and fear of acquiring infection (30). The conclusions of our work are as follows:

1. Retrieved and analyzed data revealed the following six phases of interest of SARS-CoV-2 in Polish Internet. In future we should overlap and confront them to the whole process of emergence and extinction of the pandemic. These phases could be names as: 1/ Chinese phase, 2/Italian phase, 3/ Waiting phase, 4/ Epidemic and Mitigation phase, 5/ Social distancing and Lockdown phase, 6/ Anticrisis shield, Restrictions releasing (Fig. 4).
2. We noticed the differences in platform usage as knowledge (information) providers in the following phases of epidemics. Different media (platforms) have separated informative functions. Information media (Wikipedia and Google) do not display some peaks of interest (e.g. for special act against COVID-19), because probably the awareness about the virus and the knowledge on the disease has already been saturated. After the “definition need phase 1-2” people seem to be more interested mainly in the update (news information on COVID-19) on Twitter, YouTube or other electronic media. This has covered the phase, which might be called “norms need phase 4-5”, because in this time the most searched topics were protection measures and mitigation behaviors. That is the reason that anti-crisis shield issues gathered more interest in information related media (Google, news article) as well as on politically oriented Twitter than on other social media.
3. Mitigation strategies have gathered the highest interest in all investigated platforms. What is important, dates of released declaration of measures introduction (not dates of enacting or acting in force [29]) were responsible for visible peaks of social interest. This suggests that government’s decisions were instantly filtered by social networks implying a high level of social uncertainty. This might be defined as a subtle indicator of pre-panic behaviors. This raises questions about the role of the media in managing the disease and if the ordered measures were really efficient in diminishing the pandemic scale.
4. The focus on social activity posed in this study revealed trends in common understanding of self-protective measures. This lay-referral framing system of opinions and beliefs should draw attention of public health professionals to evaluate potential epidemiological risk of infection. Our analysis showed that Poles looked for professional (or quasi-professional) measures of self-protection: “antiviral mask” or “hand disinfection” (instead simple “washing hands”). We were able to identify at least few misinformation, cognitive errors or fake news, which significantly impact interest on “Koronawirus” topic in Poland. They fall on fertile ground, due to the script mechanism of simplifying reality, where simpler explanations are more easily absorbed by people (11). Especially since epidemiologists and public health professionals do not know enough about COVID-19 yet, and recommendations and opinions of authorities such as WHO or Ministry of Health are still being updated and often being contradictory to previous opinions (e.g. effectiveness of protective mask) causing cognitive dissonance in the public. As a good example we may take the popularity of a product, which does not exist on the medical market such as an “antiviral mask” (see Appendix). We show that scientifically confirmed and effective infection control strategies such as keeping high hygienic standards are not popular among Internet users and pseudo-professional solutions are much more attractive until official recommendations are introduced (Fig. 7).
5. Low interest of the general public in COVID-19 before the disease introduction could be associated with a low epidemiological awareness of average Poles, especially when COVID-19 pandemic is massively discussed in traditional and social media by a relatively small but loud group of people (16). It’s worth to mention, that a positive effect of interest in “Koronawirus” could be a probable increase in epidemiological and hygienic knowledge in Polish population which is probably below European average (even health literacy for non-infectious diseases does not differ significantly in comparison to other European countries (21).
6. Last conclusion is rather an *ad hoc* recommendation. It seems that the Ministry of Health’s Public Relation Team should concentrated more on traditional media and get a quick response instead of using Twitter in a wrong way (e.g. providing daily updates on COVID-19 surveillance on Twitter only without any official database, which can be automatically collected by epidemiologists). Thus, due to the known and empirically confirmed during current outbreak in Poland (31) mechanism of stability seeking in the general population, the authority of the Minister of Health could be better communicated in risk perception management (32).

In addition, Internet media analysis could fill gaps in socio-medical research on collective actions during an important public health disruption event such as epidemics of infectious diseases (10, 17).

Acknowledgments: We thank PNFN (2019-21) and FU Berlin (FU AvH: 08166500) for partial financial support. We thank Łukasz Krzowski, Mariusz Duplaga, Daniel Płatek, Ireneusz Skawina, Andrzej Buda, and Marcus Doherr for fruitful discussions. Code and data are available at https://github.com/ajarynowski/koronawirus.

## Data Availability

Time series and code of analysis available in R

https://github.com/ajarynowski/koronawirus

## APPENDIX

The most important outcome shows the intensity of seeking the self-protective measures (Fig. A1, A2). On Figure A1 we can see that the amount of queries regarding protective masks reached around 50 thousand queries daily. The most interesting got: “protective mask”. This is a very interesting fact, because before the introduction of SARS-CoV-2 in Poland (before 4.03.20) polish health minister Łukasz Szumowski and other health professionals did not recommend masks as individual protection measures. The line on Figure A1 shows that Internet users seeked information before the first case of coronavirus and also when masks were recommended and stated as the condition of movement in public areas.

**Figure A1.**
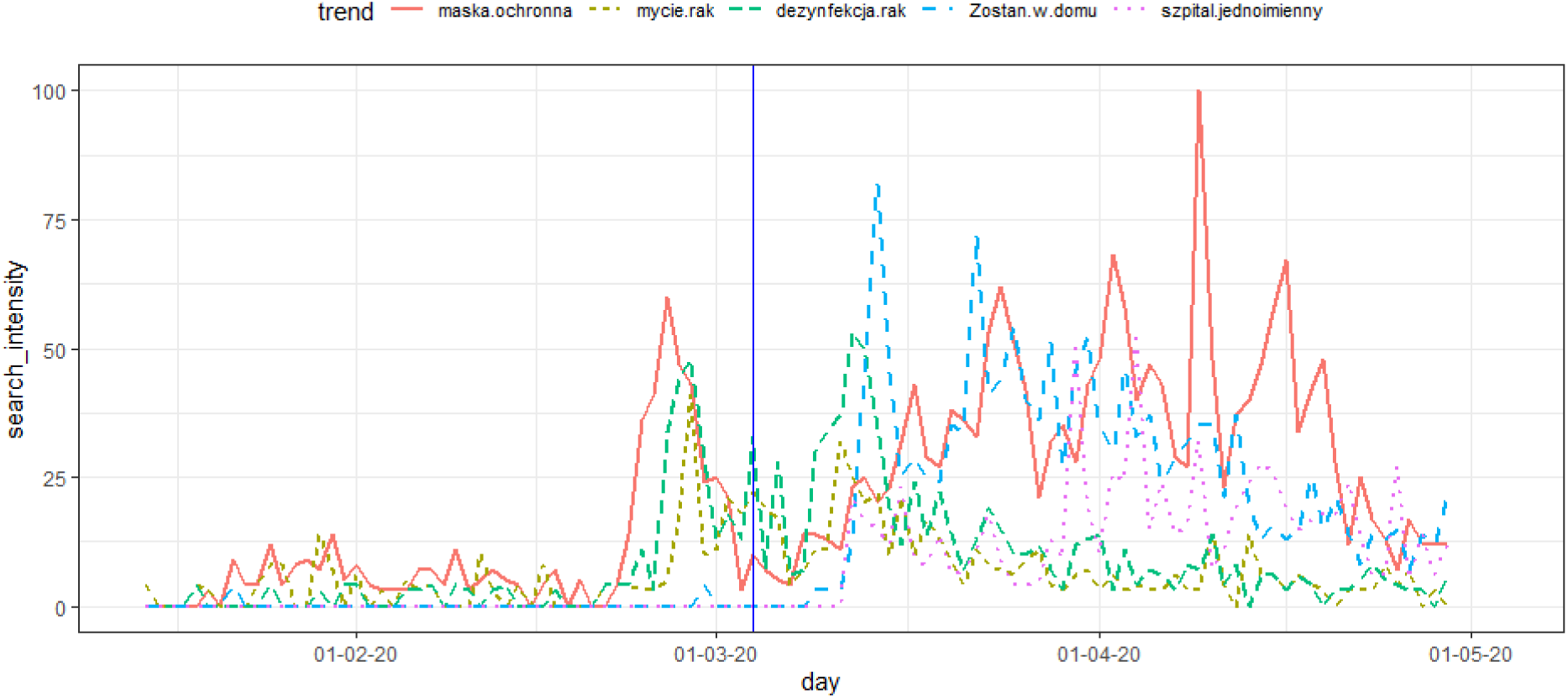
The intensity of queries with the phrases “protective mask (big)”, “hand washing”, “hand disinfection”, “Stay at Home”, “COVID-19 hospital” in Polish Google (15.01-07.04.20) generated using the Google Trend tool. Disease introduction marked with the vertical line.

**Figure A2.**
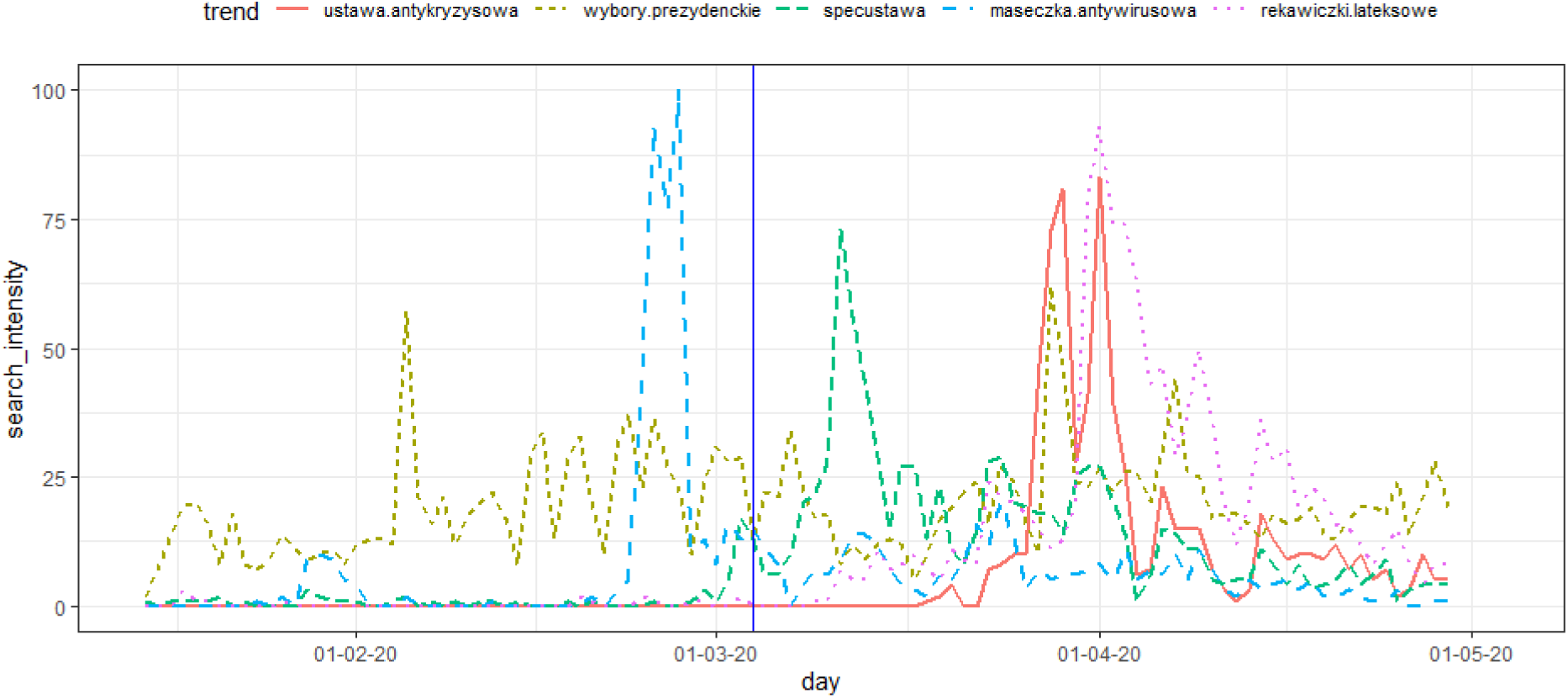
The intensity of queries with the phrases “anticrisis law”, “presidential election”, “special act on COVID-19”, “antiviral mask”, “latex gloves” in Polish Google (15.01-29.04.20) generated using the Google Trend tool. Disease introduction marked with the vertical line.

People were searching for information on further epidemiological topics related to infection and protective measures as well: “hand washing”, “hand disinfection”, “Stay at Home”, “COVID-19 hospital” (Fig. A1). The most popular epidemiological queries as latex gloves reached around 100 thousands queries daily (Fig. A2) (own estimates based on Google Ads) in peak. It should be noted that professional vocabulary such as “hand hygiene” practically does not appear in queries (below the “noise threshold” compared to other epidemiological terms (Fig. A1, A2)). Queries related to hand hygiene had a peak in late February and beginning of March. Queries related to masks had their peak of popularity at the end of February and the lack of increase in popularity in March (compared to other epidemiological queries (Fig. A1, A2)) may be due to the success of information campaigns on their alleged low effectiveness (in protecting users) or simply due to the lack of the masks on the market.

**Figure A3.**
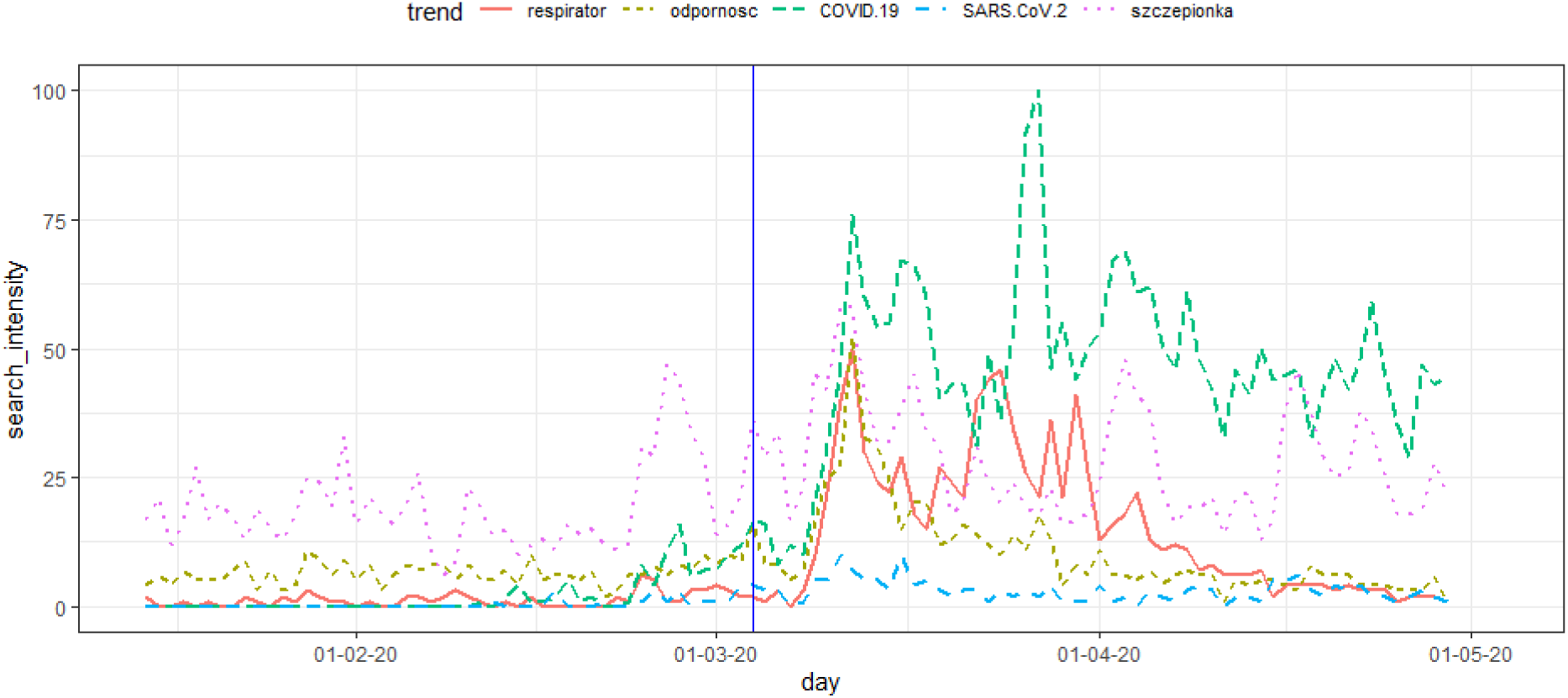
The intensity of queries with the phrases “respirator”, immunity”, “COVID-19”, “SARS-CoV-2”, “vaccine” in Polish Google (15.01-29.04.20) generated using the Google Trend tool. Disease introduction marked with the vertical line.

Medical queries as ‘respirator’ and ‘immunity’ can reach around 20 thousands queries daily (Fig. A3) (own estimates based on Google Ads) in peak. However the most popular terms are related to political issues as anticrisis shield, which can reach around 200 thousands queries daily (Fig. A4) (own estimates based on Google Ads) in peak. There is a controversial issue of presidential elections, which are supposed to take place on 10.05.20 (Fig. A2). Moreover, Minister of Health - Łukasz Szumowski is also intensively searched in Google (Fig. A4), and he became on 07.04.20 the most trusted person in Poland (S1).

**Figure A4.**
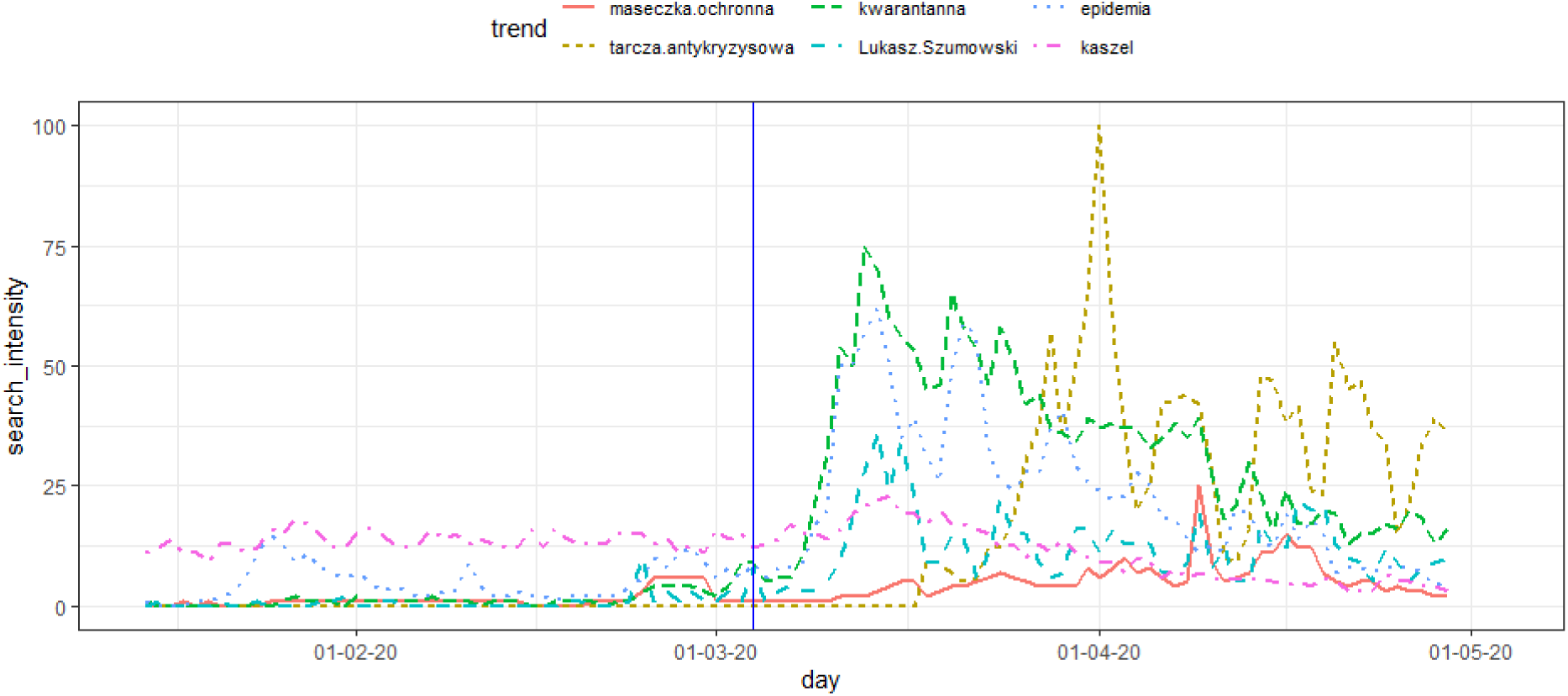
The intensity of queries with the phrases “protective mask”, “anticrisis shield”, quarantine”, “minister of health”, “epidemic”, “cough” in Polish Google (15.01-29.04.20) generated using the Google Trend tool. Disease introduction marked with the vertical line.

We also checked symptomatic queries such as fever, cough (Fig. A4), shortness of breath and anosmia, etc., but they do not vary significantly in time. However, this analysis may be repeated (S2) if a higher burden of disease will be the case.

**Figure A5.**
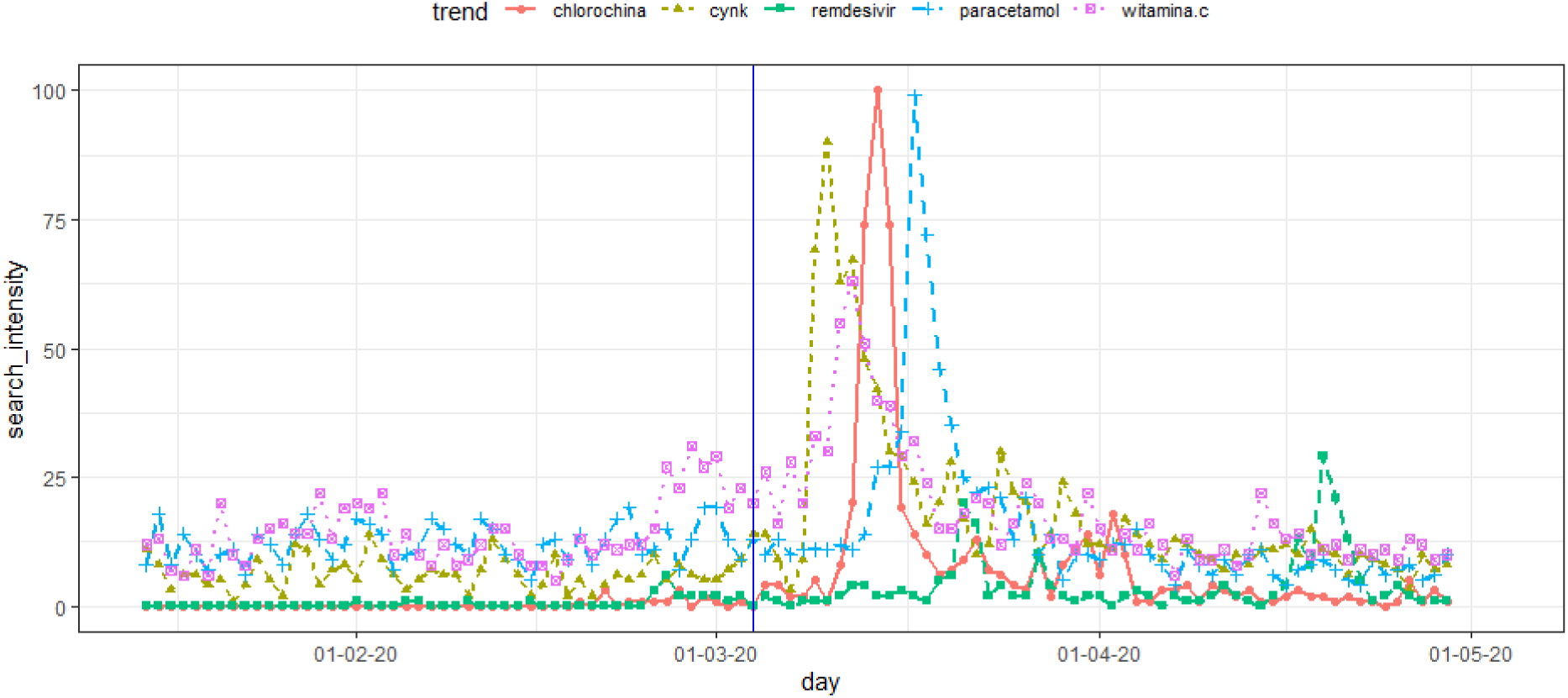
The intensity of queries with the phrases “Chloroquine”, Zync”, “Remdesivir”, “Paracetamol”, “vitamin C” in Polish Google (15.01-29.04.20) generated using the Google Trend tool. Disease introduction marked with the vertical line.

Pharmaceutical queries as Chloroquine can reach around 50 thousands queries daily (Fig. A5) (own estimates based on Google Ads) in peak.

S1. IBRIS. Łukasz Szumowski nowym liderem rankingu zaufania; 2020. Accessed: 2020-04-13. https://wiadomosci.onet.pl/tylko-w-onecie/koronawirus-sondaz-lukasz-szumowski-politykiem-z-najwiekszym-zaufaniem/.
S1. Times. Google Searches Can Help Us Find Emerging Covid-19 Outbreaks; 2020. Accessed: 2020-04-10. https://www.nytimes.com/2020/04/05/opinion/coronavirus-google-searches.html.

